# Factors associated with utilization of preconception Sickle cell screening among women receiving antenatal care at Hoima regional referral hospital, Uganda

**DOI:** 10.1101/2024.05.07.24306985

**Authors:** Solomon Kyakuha, Mbabazi G. Scovia, Nathan Mugenyi, Andrew Twineamatsiko

## Abstract

**Introduction:** Globally, it is estimated that over 300,000 babies are born with Sickle cell disease annually, yet the uptake of preconception sickle cell screening services is at 25% among the total population that become pregnant every year. In Uganda, about 20,000 babies are born annually but with a low utilization rate of Pre-Conception Sickle cell screening services among women at 11%. This study aimed at determining the utilization rate and factors associated with preconception sickle cell screening among pregnant women attending antenatal care at Hoima regional referral hospital, Uganda.

**Methodology:** A hospital-based cross-section study was done, systematic sampling used, and participants subjected to a semi-structured questionnaire. 334 participants were recruited, and data was collected, entered into Epidata, and analyzed using STATA version 14.

**Study findings:** The utilization rate of Pre-Conception Sickle cell screening services among women receiving Ante Natal Care was at 11.4%. There was a significant association between woman’s level of education, marital status, partners support and knowledge towards Pre-Conception Sickle cell screening services.

**Conclusions:** The utilization rate of Pre-Conception Sickle cell screening services is low with level of education, marital status, and having familial history of sickle cell disease, partner support and awareness being significant associated factors.

**Recommendation:** There is need to empower women to attain a formal education through adult literacy programs, massive sensitization and conducting regular health talks regarding Pre-Conception Sickle cell screening services.

## Introduction

Sickle cell disease (SCD) is a parasol term for several abnormal hemoglobin molecules categorized by the presence of beta-globin gene mutations in which at least one of the point mutations leads to production of hemoglobin S. Sickle cell disease is associated with a variety of complications, which range from hemolytic anemia, chronic end organ damages and premature death. It’s noted that in developed countries such as United States, about 95% of the children with sickle cell disease grow to adulthood that is above eighteen years compared to 84% in Uganda (1), thus resulting in affected adults with significant co-morbidities, complex medical complications. Important to note, there are a few sickles cell specialists in Uganda and most of these few are situated in the urban areas leaving the rural with no option but get care from none specialists, thus decreasing the quality of life of those children with the disease (1).

There is a rise in complications among those with SCD such as high death rates, and social and economic stress. Parents of children diagnosed with sickle cell disease suffer a lot of emotional distress and guilt in Africa. This is associated or attributed to low knowledge levels among parents, and they end up attributing the disease to socio-cultural, religious and economic scopes. To the worst extend, sickle cell disease has been thought to be a curse in the family thus causing a lot of stigma and expensive medical costs. This has left some families divorced leading to single parenthood (2).

Globally, it is estimated that over 300,000 babies are born with SCD, and majority of these cases arise from Sub Saharan Africa, Nigeria leading with a total of 150000 children. SCD is frequently lethal in the early life and is responsible for 70-80% killings of afflicted children before the 5th year of age in sub-Saharan Africa. Sickle cell anemia is responsible for 20% of the annual childhood deaths among children with SCD in Uganda (3).

PCSCS is a technique carried out among partners (potential parents) before conception to screen a collection of hemoglobinopathies characterized by a 2-beta errors resulting in production of hemoglobin S. Failure to screen results in delivery of children with either sickle cell disease (SCD) or sickle cell traits (SCT) also known as carriers (4).

Counseling and hereditary screening services are present in many facilities and institutions in many cities of many countries; however, preconception and premarital screening and counseling of genetic disorders offered are limited.

Preconception counseling and testing have been thought about as one of the strategies to reduce sickle cell burden as evidenced in areas of high endemicity such as Cyrus and Sardinia. Preconception care is much understood by communities as a pre-pregnancy care and which is mostly considered for women, and this is misleading, and yet it should involve both male and female. Preconception care involves services other than preconception sickle cell screening such as family planning services, infertility information, HIV testing and treatment, hepatitis B testing and treatment and many others.

Preconception sickle cell screening provides several advantages with the key main ones being to determine whether the parents to be either carriers of genetic condition (SCD) and basing on results of the screening or testing expert counseling is done to the prospective parents to assist them in making informed decisions as they plan for becoming pregnant (Carrier Screening, 2018).

Research has shown that despite couples knowing their status, 88% of the tested couples will go on to bare children, but have an early intervention to have their children screened for SCD (6).

Globally efforts of primary prevention of sickle cell disease range from legal mandates of preconception hemoglobinopathy screening to public education and awareness. Previously preconception screening for SCD was thought to decrease the sickle cell burden majorly in areas where marriage was organized customary; however, changing cultural norms and preferences for marriage have hindered the success of the program in the current era (7).

Globally, it is estimated that the uptake of preconception sickle cell screening services is at 25% among the total population that become pregnant every year. This uptake is proven to decrease the burden of SCD among children by only 4%, thus if the uptake is increased beyond that, this will result in a more decrease in the burden (8). In sub Saharan Africa the uptake of PCSCS is estimated at 6% according to the study done at Paul Moukambi regional hospital Centre in koula-moutou, this is much lower compared to the uptake globally among child bearing age (9). In East Africa, many of the parents of children with sickle cell get to know their statuses after diagnosis of SCD among their children. There was an expression of participants to make preconception screening obligatory (10).

In 2018, the royal decree of Saudi Arabia mandated the preconception screening of beta thalassemia and SCD before drafting marriage. Studies done in the United States, Jamaica showed similar results that despite an increased knowledge on SCD, this doesn’t affect the decision of marriage choice. In Saudi Arabia, of the 934 participants, it was noted that 88% of the participants with sickle cell trait still chose to marry each other. After knowing their status couple is well prepared for the outcome in their children such as early diagnosis and treatment of sickle cell complications such as early initiation on folic acid and urea (11).

In another cross-sectional study done in south Saudi Arabia among women who had done preconception sickle cell screening and their partners, a total of 413 participants were subjected to online questionnaire and it was established that 66.8% of the participants had similar sickle cell status results, 67.8% were carriers for SCD and of the 413, only 46.5% had attended preconception counseling clinics, over 50% rejected the counseling after sickle cell testing. It was noted that 18.2% of the participants had proceeded into marriage because they thought disease transmission to children was small and 5.2% proceeded because they thought that the lives of their children wouldn’t be affected by disease. From the study it was noted that cultural, religious, and financial factors greatly affect the preconception sickle cell screening, with culture contributing about 48% (7).

In a study conducted among Dutch Moroccan and Turkish married women, it was noted that four key factors are associated with their perception towards preconception screening, these included the choice of their partners to have a sickle cell test done, the choice of marriage that is if one decides to marry from a population with high prevalence of sickle cell disease and choice of having a healthy child free of sickle cell disease. Other factors that influenced their perspective towards preconception sickle cell screening included their religious affiliations, culture, and gender (12).

In another study done in Benue state Nigeria, it assessed factors associated with awareness of hemoglobin statuses among pregnant women, these factors included age, type of occupation, education levels, level of income, reception of antenatal care, number of pregnancies, and distance from the health facility. From the study it was noted that younger participants had higher chances of being aware of their status compared to older ones, comparing the unemployed, farmers were 5.8 times lower odds whereas traders had 1.2 times higher odds of being aware of their statuses. The odds of mothers attending antenatal care was 2 times higher compared to those who didn’t attend (13).

According to study done in Uganda by Ndeezi et al in 2016 to determine the prevalence of sickle cell trait and disease, samples were collected from 112 districts across the country, a total of 97,631 results of the 99,243 samples were available. The overall children with sickle cell trait were 13.3% and those with disease were 0.7%. Different regions had different numbers of SCD and sickle cell trait (SCT). It was noted that SCD was 0.2% in southwestern and 1.5% in east central while SCT was 4.6% in southwestern (Kabale, Kisoro) and 19.8% in east central (Buikwe). The lowest prevalence of 3.0% was seen in two districts and eight districts had prevalence of above 20%, with the biggest being 23.9% in Alebtong (14).

In another study done in Luuka district to assess the readiness for implementation of preconception care in Uganda. Preconception care guidelines are well described in the maternal nutrition, adolescent and reproductive health policies. These guidelines have changed over time to include services such as hepatitis B screening, Rubella vaccination, management of health conditions such as hypertension, diabetes, and many others. It is noted in several studies that increasing knowledge and awareness are key in increasing preconception care services usage and attendance however there is an increased demand to disseminate preconception care packages in the district health system using best package and delivery channels. There are several barriers to preconception service utilization, such as low-income settings, Knowledge gap among mothers and low awareness levels (15).

In a study done at Mulago hospital among prim gravid mothers, its noted that despite 72.2% of mothers having positive attitude towards SCD screening, only 65.4% had adequate knowledge about preconception sickle cell screening, 49% knew it could be tested in blood.11% had screened for the disease before conception and about 66% supported the idea of making preconception screening mandatory (16).

Despite it being known that preconception sickle cell screening provides a key role in primary prevention and mothers having a positive attitude towards sickle cell screening and it being integrated into health care system, the uptake remains low. There is no study done in Hoima regional referral hospital to determine the uptake of preconception sickle cell screening and the associated factors despite having studies in Uganda, thus the study aims to establish the factors that are associated with the utilization of PCSCS among women of reproductive age.

## Methods and Materials

### Study design

The study design was a cross-sectional study. Cross-sectional study design was used at one point in time and there was no follow up of participants after the study. It’s a recommended measure of association and allowed several participants to be recruited at the same time.

### Study site

The study site was HRRH, located about 200km west of Kampala city. It had total bed capacity of 360 beds and served as a regional hospital for eight districts namely, Hoima, Kakumiro, Kagadi, Kibaale, Buliisa, Masindi, Kiryandongo and Kikuube. The hospital runs both inpatient, and outpatient services. It provides both specialized and non-specialized health care services. The hospital serves about 500,000 patients annually and 80 women for antenatal care services per day. This study was carried out at maternal and child health clinic, in the department of obstetrics and gynecology. Study was carried out at Hoima regional referral hospital because there were no similar studies carried out in the region and only available information was from central, western and eastern part of the country.

### Study population

All pregnant women in their reproductive age 18 to 49 years, attended antenatal care at HRRH.

### Sample population

The sample population included all pregnant women aged 18 to 49 years. Due to ethical considerations, those below 18 years were regarded as minors and couldn’t independently consent to participate in the study.

### Sampling frame

This included a list of all pregnant women aged 18 to 49 years who had registered to receive antenatal services at HRRH.

### Sampling techniques

Participants would be obtained using systematic probability sampling. This was to ensure that each mother had an equal chance to participate in the study (Saunders, Lewis, & Thornhill, 2003)

### Sampling procedure

Every fourth mother in the daily ANC register would be selected. The sampling interval was obtained by using the formula below.

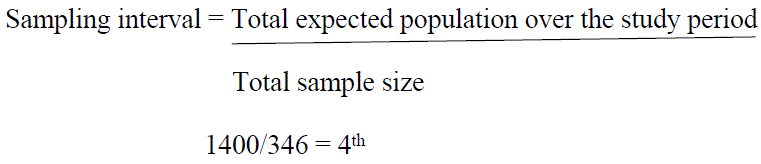

The first participant was obtained by simple random sampling (one of the first three participants) to avoid bias.

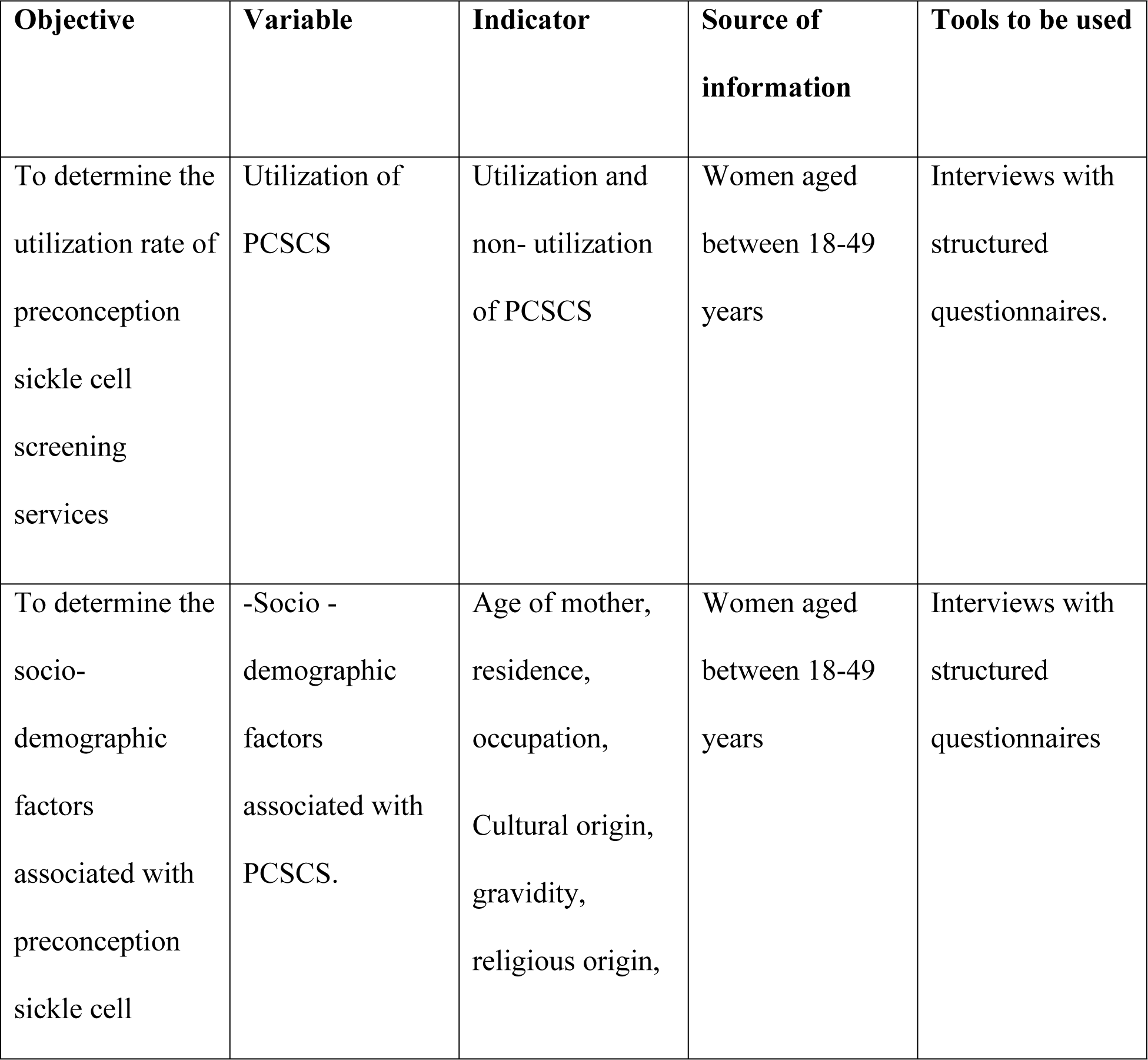
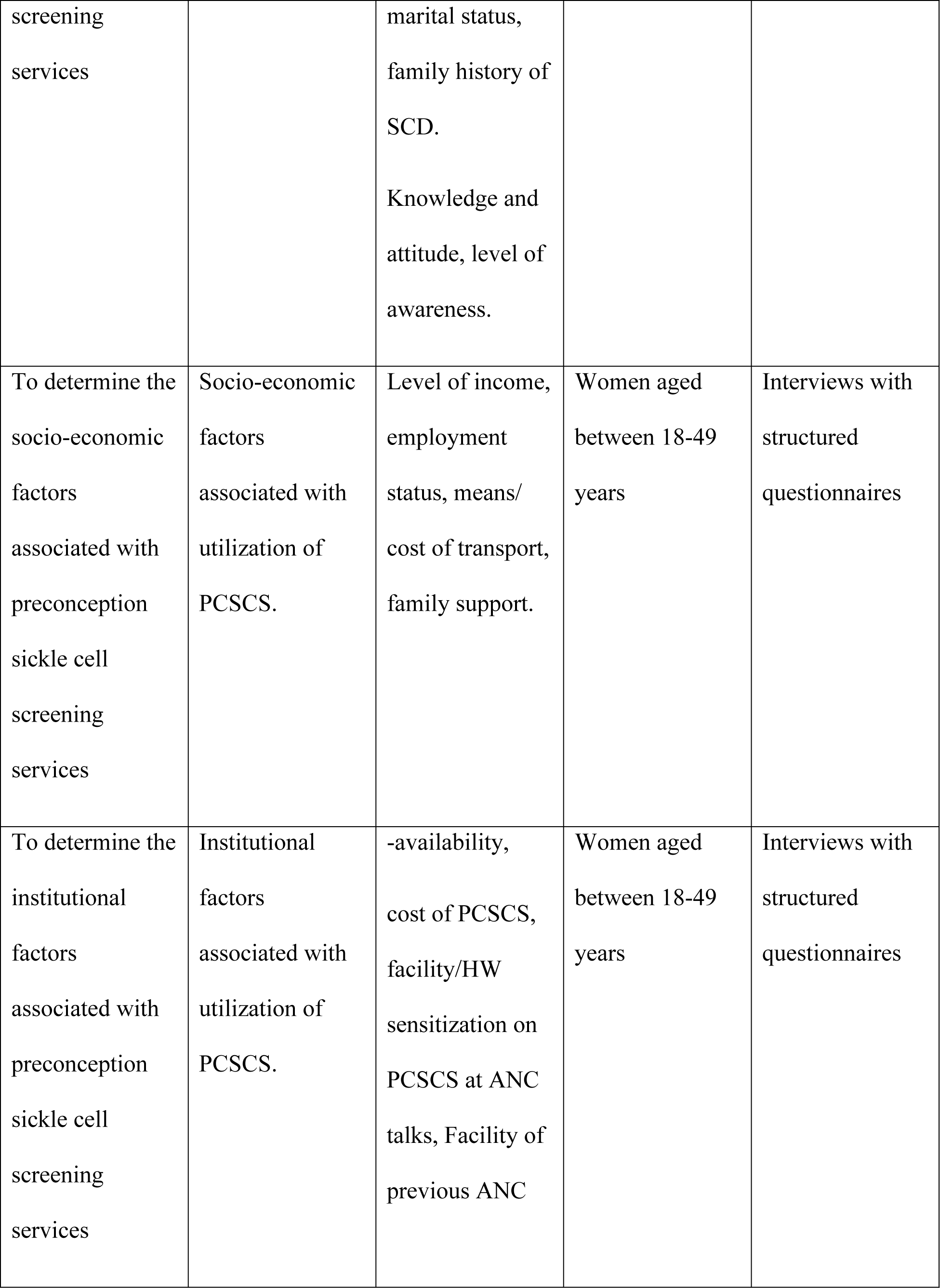
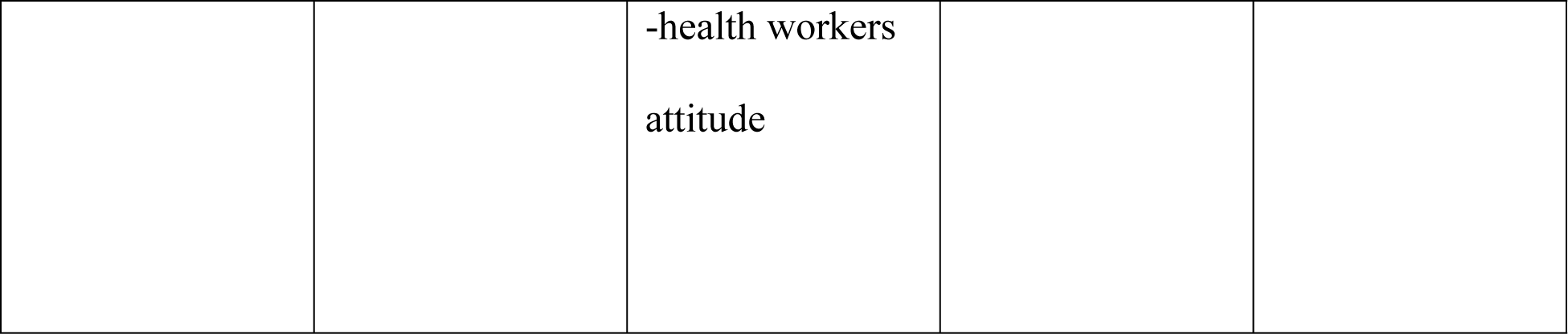
Table of variables.

**Table of variables**
| Objective | Variable | Indicator | Source of information | Tools to be used |
| --- | --- | --- | --- | --- |
| To determine the utilization rate of preconception sickle cell screening services | Utilization of PCSCS | Utilization and non- utilization of PCSCS | Women aged between 18-49 years | Interviews with structured questionnaires. |
| To determine the socio-demographic factors associated with preconception sickle cell | -Socio - demographic factors associated with PCSCS. | Age of mother, residence, occupation, Cultural origin, gravidity, religious origin, | Women aged between 18-49 years | Interviews with structured questionnaires |
| screening services |  | marital status,<br>family history of SCD.<br>Knowledge and attitude, level of awareness. |  |  |
| To determine the socio-economic factors associated with preconception sickle cell screening services | Socio-economic factors associated with utilization of PCSCS. | Level of income, employment status, means/ cost of transport, family support. | Women aged between 18-49 years | Interviews with structured questionnaires |
| To determine the institutional factors associated with preconception sickle cell screening services | Institutional factors associated with utilization of PCSCS. | -availability, cost of PCSCS, facility/HW sensitization on PCSCS at ANC talks, Facility of previous ANC | Women aged between 18-49 years | Interviews with structured questionnaires |
|  |  | -health workers<br>attitude |  |  |

## Data collection methods

Data was collected using a face-to-face interview with open and close-ended questions about utilization of PCSCS services and its associated factors. This was used because it could easily provide the information needed from participants to measure PCSCS utilization, and it could cover a large number of respondents in the least time possible.

## Data collection tools

A semi-structured questionnaire was used to collect information about utilization of PCSCS and the factors associated with its utilization. The questionnaire was administered by the researcher to the respondent by use of a face to face interview.

## Ethical considerations

The study received ethical clearance from Uganda Martyrs University, Mild May Uganda Research Ethics Committee (MUREC) and HRRH research committee, since it involved highly sensitive information to be collected. Participants were not identified by their names or contacts or address to promote participant’s privacy. For this study in particular, I also thought approval from Hoima RRH management to allow me carry out the study at the site.

## Data Analysis Plan

Descriptive statistics was used to summarize continuous variables into mean and standard deviations or median and interquartile ranges, while frequencies and percentages will be used to summarize categorical variables. A 95% confidence interval was used for utilization of PCSCS. Pearson’s Chi square test was carried out to establish the factors associated with utilization of PCSCS services at a 5% level of significance using STATA version 14.

## Data quality control

The research principle investigator trained three research assistants for two days on how to collect data. The research assistants double-checked all the questionnaires for completeness after the participants had finished responding to the questionnaire. The questionnaires were pretested using five participants before the study. All data entered into Epidata was to be password protected and backed up in Google drive and the questionnaires would be kept under lock and key drawers.

## Results

Of the total estimated sample population (346 participants), the study only involved 333, the 11 participants declined to participate, 10 participants withdrew from the participation during the interview process. This gave a 96% respondent rate. The study results were presented in accordance to the study objectives as follows:

## Utilization rate of preconception sickle cell screening services among women of reproductive age

In accordance to the utilization of preconception sickle cell screening services among the 334 participants (women), the utilization rate of PCSCS was at 11.4%(38), 95%CI (8.0-14.8) of women aged 18-49 years that were receiving antenatal care from antenatal clinic of HRRH at the time of study and 88.6%(296),95% CI(85.2-92.0) had not utilized the preconception sickle cell screening services as illustrated in the figure 1 below.

**Figure 1.**
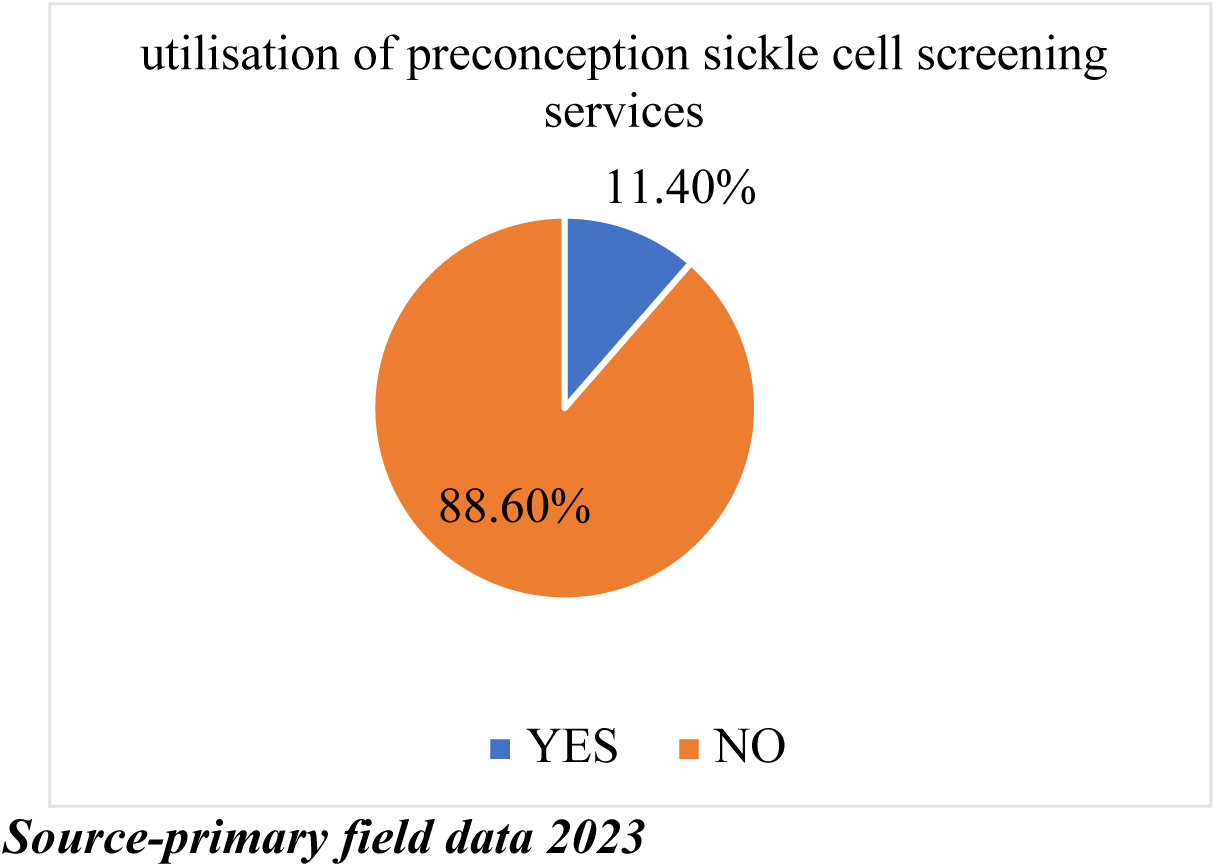
**Shows a pie chart indicating the percentage of women aged between 18-49years who had utilized PCSCS at HRRH**

## Socio-demographic characteristics of women 18-49 years receiving antenatal care from H RRH

Respondents who received antenatal care from Hoima regional referral during the study period were asked about the age, marital status, place of residence, employment status, and history of sickle cell disease, religious affiliations, and level of education, tribe and number of children.

From the study, it was noted that respondents were distributed across all age groups with majority of the participants were between ages of 18-35 years of age (98.7%) and the rest were between 36-49 years (1.3%). Also majority of the respondents were cohabiting with their male partners 141(42.3%), followed by those who were married 99(28.8%), singles 27.6(27.6%) and least by those who were windowed by the time of the study 4(1.2%).

Furthermore, it was found that 165(49.6%) of the respondents who participated in the study had completed secondary education, 99(29.7%) primary, 46(13.8%) tertiary (diploma), 13(3.9%) university(degree), 10(3%) uneducated (no formal education). This implied that majority had completed secondary and only 3% had not attended any formal education.

For employment status, majority of the respondents were employed 216 (64.7%) and 118(35.3%) were unemployed while for area of residence, majority of the respondents were living in the urban areas 223(69.3%), followed by those in rural 76(23.6%) and minority of them were residing in both urban and rural areas 23(7.1%) at the time of the study or time when data was being collected.

The religious affiliations of the respondents were grouped into five major categories, catholic, Anglican, Pentecost, Muslim and others. Of the total number of respondents, majority were Catholics 140(41.9%), followed by Anglicans 127(38.0%), then Pentecost and Moslem with equal numbers 29(8.7%) and last others with 9(2.7%). The other religious affiliations included seventh day Adventist, faith of unity, earthiest and the non-believers.

It was noted that over 69.5 %(232) of the respondents were already living with at least one child and more and about 102(30.5 %) were first time mothers (prime gravid) receiving antenatal care from HRRH during the period of data collection.

During the study, the tribal origin of the respondents was divided into five major tribes common in the study area that is Banyoro, Baganda, Bakonjo, Bakiga and others. It was found out that majority of the respondents were Banyoro 216(64.9%), other tribes (Batoro, Barulu, Basoga and many others) 67(20.1%), Baganda 29(8.7%), Bakiga 20(6%) and Bakonjo 1(0.3%).

Of the total number of respondents(334), it was found out that 51(15.3%) of them had a familial history of sickle cell disease from either first or second degree relatives on either maternal or paternal sides and majority of the respondents 283(84.7%) did not report any history of sickle cell disease.

Finally, it was noted also that over 123(36.9%) were earning between 100,000 to 1million, 120(36%) were earning below 100000, 84(25.2%) were not earning anything and only 6(1.8%) earned above 1 million every months. There were different sources of these earnings which included, farming, small business enterprises such as saloons, poultry and salary from formal employment such as nurses, teachers. Some women reported that some earnings were provided by their husbands as stipend.

## Socio-demographic factors associated with PCSCS among women18-49 years receiving ANC at HRRH

From the table 2 above, several socio-demographic factors were analyzed for their relationship or association with utilisation preconception sickle cell screening services among women 18-49 years that were receiving antenatal care from Hoima regional referral hospital. These included age, education level, employment status, residence, tribe, marital status, religion and history of sickle cell.

**Table 2.** Socio-demographic characteristics of the women aged 18-49years receiving ANC at HRRH.

| Variable (n=334) | Frequency | Percentage |
| --- | --- | --- |
| **Age in years** |  |  |
| 18-35 | 328 | 98.7 |
| 36-49 | 6 | 1.3 |
| **Marital Status \*** |  |  |
| Married | 96 | 28.8 |
| Single | 92 | 27.6 |
| Cohabiting | 141 | 42.3 |
| Widowed | 4 | 1.2 |
| **Education level\*** |  |  |
| Uneducated | 10 | 3.0 |
| Primary | 99 | 29.7 |
| Secondary | 165 | 49.6 |
| Diploma | 46 | 13.8 |
| Degree | 13 | 3.9 |
| **Employment status** |  |  |
| Yes | 216 | 64.7 |
| No | 118 | 35.3 |
| **Residence\*\*** |  |  |
| Urban | 223 | 69.3 |
| Rural | 76 | 23.6 |
| Both | 23 | 7.1 |
| **Religion** |  |  |
| Catholic | 140 | 41.9 |
| Anglican | 127 | 38.0 |
| Pentecost | 29 | 8.7 |
| Moslem | 29 | 8.7 |
| Others | 9 | 2.7 |
| **Have children** |  |  |
| Yes | 232 | 69.5 |
| No | 102 | 30.5 |
| **Tribe\*** |  |  |
| Munyoro | 216 | 64.9 |
| Muganda | 29 | 8.7 |
| Mukonjo | 1 | 0.3 |
| Mukiga | 20 | 6.0 |
| Others | 67 | 20.1 |
| **SCD history** |  |  |
| Yes | 5 | 15.3 |
| No | 283 | 84.7 |
| **Monthly income\*** |  |  |
| Below 100,000 | 120 | 36.0 |
| 100000 to 1 Million | 123 | 36.9 |
| Above 1 Million | 6 | 1.8 |
| None | 84 | 25.2 |

Total number of participants was 334, \*total number of participants 333
\*\*total number of participants was 322

**Table 3:** Bivariate analysis of Socio-demographic factors associated with PCSCS among women18-49 years receiving ANC at HRRH.

| **Variable** | **PCSCS**<br>**Utilization**<br>**Yes n (%)** | **No n (%)** | **Chi-Square** | **P-value** |
| --- | --- | --- | --- | --- |
| **Age** |  |  | 3.48 | 0.169 |
| 18-35 | 37(11.9) | 275(88.1) |  |  |
| 36-49 | 1(4.8) | 20(95.2) |  |  |
| **Education level** |  |  |  |  |
| Uneducated | 0(0.0) | 10(100) | 3.64 | 0.484 |
| Primary | 10(10.1) | 89(89.9) |  |  |
| Secondary | 18(10.9) | 147(89.1) |  |  |
| Diploma | 6(13.3) | 39(86.7) |  |  |
| Degree | 3(23.1) | 10(76.9) |  |  |
| **Employment** |  |  |  |  |
| **Status** |  |  | 2.67 | 0.102 |
| Employed | 20(9.3) | 195(90.7) |  |  |
| Unemployed | 18(15.3) | 100(84.8) |  |  |
| **Marital status** |  |  | 4.70 | 0.195 |
| Married | 16(16.8) | 79(83.2) |  |  |
| Single | 8(8.7) | 84(91.3) |  |  |
| Cohabiting | 13(9.2) | 128(90.8) |  |  |
| Widowed | 0(0.0) | 4(100) |  |  |
| **Family History of** |  |  |  |  |
| **SCD** |  |  | 8.75 | 0.003* |
| Yes | 12(23.5) | 26(9.2) |  |  |
| No | 39(76.5) | 256(90.8) |  |  |
| **Religion** |  |  | 3.97 | 0.410 |
| Catholic | 14(10.0) | 126(90.0) |  |  |
| Anglican | 15(11.9) | 111(88.1) |  |  |
| Pentecost | 3(10.3) | 26(89.1) |  |  |
| Moslem | 6(20.7) | 23(79.3) |  |  |
| others | 4(0.0) | 9(100) |  |  |
| **Residence** |  |  | 1.19 | 0.551 |
| Urban | 24(10.8) | 199(89.2) |  |  |
| Rural | 7(9.3) | 68(90.7) |  |  |
| Both | 4(17.4) | 19(82.6) |  |  |
| **Tribe** |  |  | 0.95 | 0.917 |
| Banyoro | 25(11.6) | 190(88.4) |  |  |
| Baganda | 4(13.8) | 25(86.2) |  |  |
| Bakonjo | 0(0.0) | 1(100) |  |  |
| Bakiga | 3(15.0) | 17(85.0) |  |  |
| Others | 6(9.0) | 61(91.0) |  |  |

*Source primary filed data 2023\* statistically significant at $P < 0.05$*

History of SCD in the family, the utilization of PCSCS was highest among those who had no familial history of SCD 39(76.5%) compared to those who had history of SCD 12(23.5%). And for p-value of 0.035 less than 0.05, there was significant relationship between history of SCD and utilization of PCSCS among women receiving ANC from HRRH.

Important to note, all the other socio-demographic factors had no significant association with utilization of PCSCS among women receiving ANC from HRRH.

## Socio-economic factors associated with utilization of PCSCS among women receiving ANC from HRRH

**Table 4:**
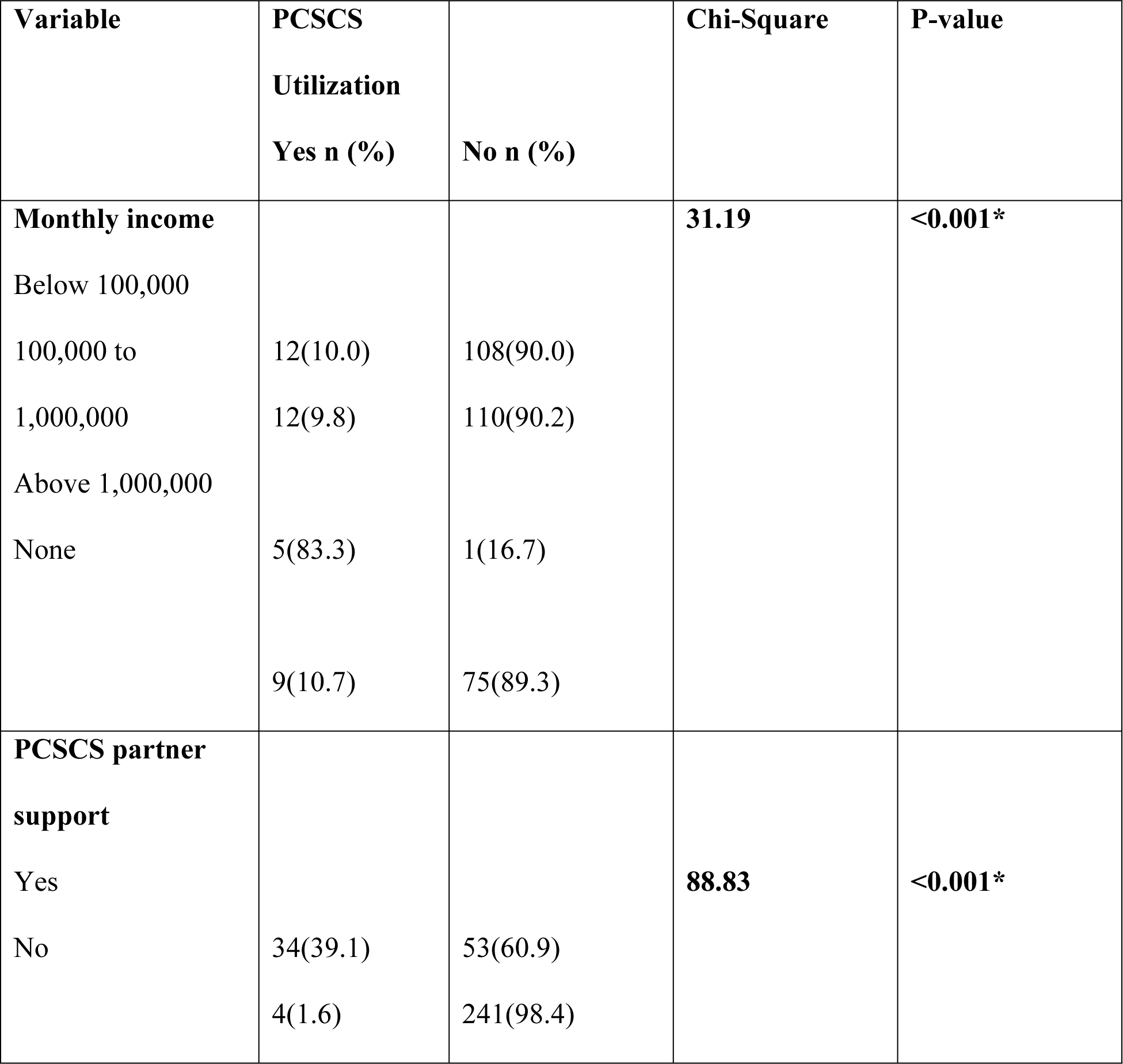
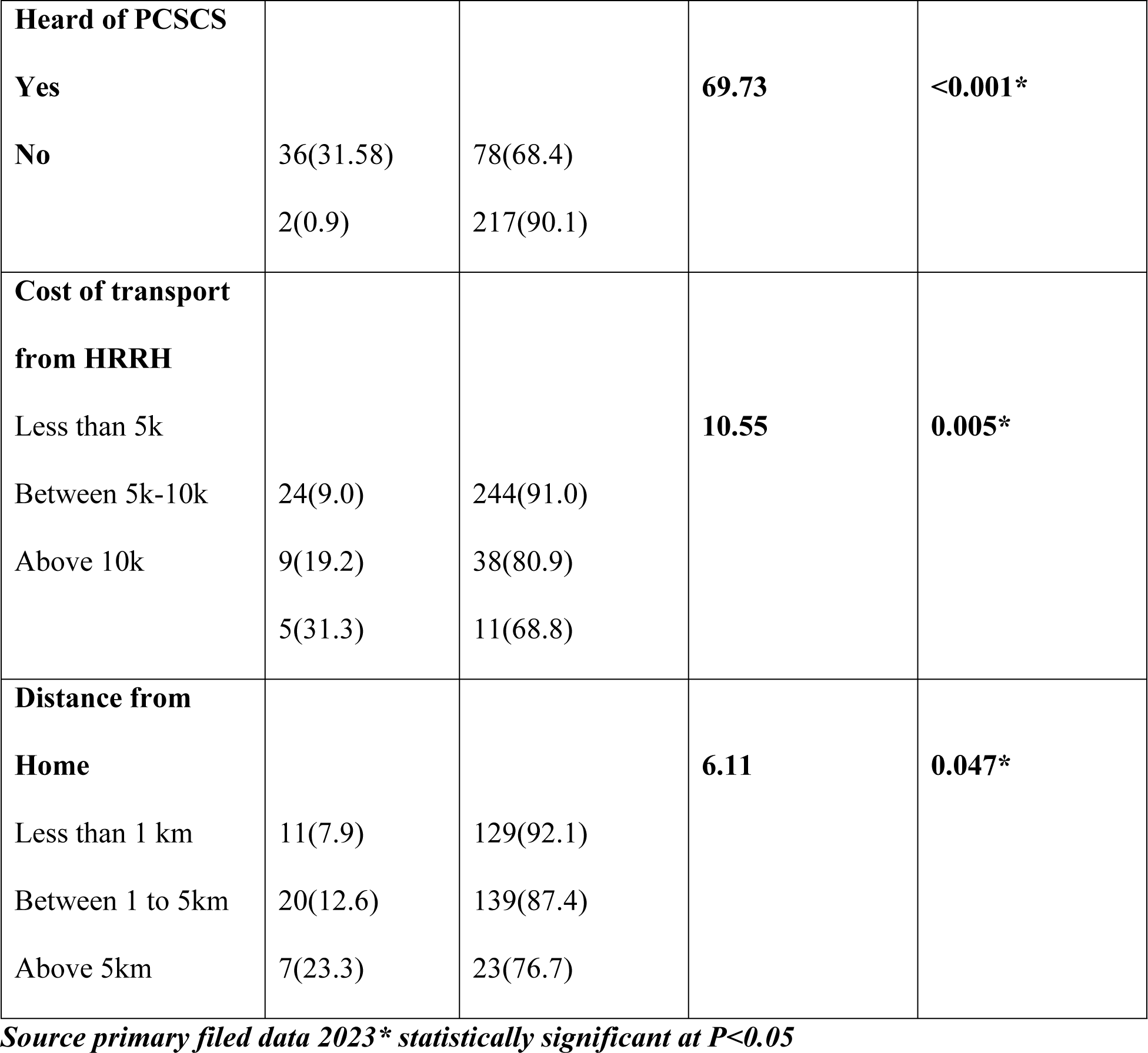
Bivariate analysis for Socio-economic factors associated with utilization of PCSCS among women 18-49 years receiving ANC from HRRH.

For the women’s monthly level of income, Utilization of PCSCS was a highest among those earned above 1000000 5(83.3%), followed by those who didn’t earn anything 9(10.7%), then those below 100000 12(10.0%) and least by those between 100000-1000000 12(9.8%). For P=0.001, there is significant association between level of income of women 18-49 years and utilization of PCSCS.

Secondary, utilization of PCSCS was highest among women who had previously heard about PCSCS from the different sources, that is media, health facility, churches 36(31.58%) compared to those who had never heard about PCSCS 2(0.9%). For the P-0.001, there is a significant association between women having heard about PCSCS and utilization of PSCS.

Thirdly, there was an increased utilization of PCSCS among partners who had partners support towards having PCSCS 34(39.1%) compared to those women who didn’t have a partner’s support 4(1.6%). And the P-0.001, there was a significant association between male partner support and utilization of PCSCS among women aged 18-49 years.

For distance of women receiving ANC from HRRH, utilization of PCSCS was highest among those who travel above 5km 7(23.3%), followed by those who travel between 1-5km 20(12.6%) and least among those who travel for less than 1km 11(7.9%). And the p-0.047, there was significant relationship between distance from HRRH and utilization of PCSCS among women 18-49 years receiving ANC from HRRH.

For cost of transport, utilization of PCSCS was highest among those who paid transport above 10000 5(31.3%), followed by those whose cost of transport was between 5000-10000 9(19.2%) and least among those who paid transport less than 5000 24(9.0%). The P-0.05 significant relationship between cost of transport and utilization of PCSCS among women 18-49 years receiving ANC from HRRH

## Institutional factors associated with utilization of PCSCS among women 18-49 years receiving ANC from HRRH

**Table 5:**
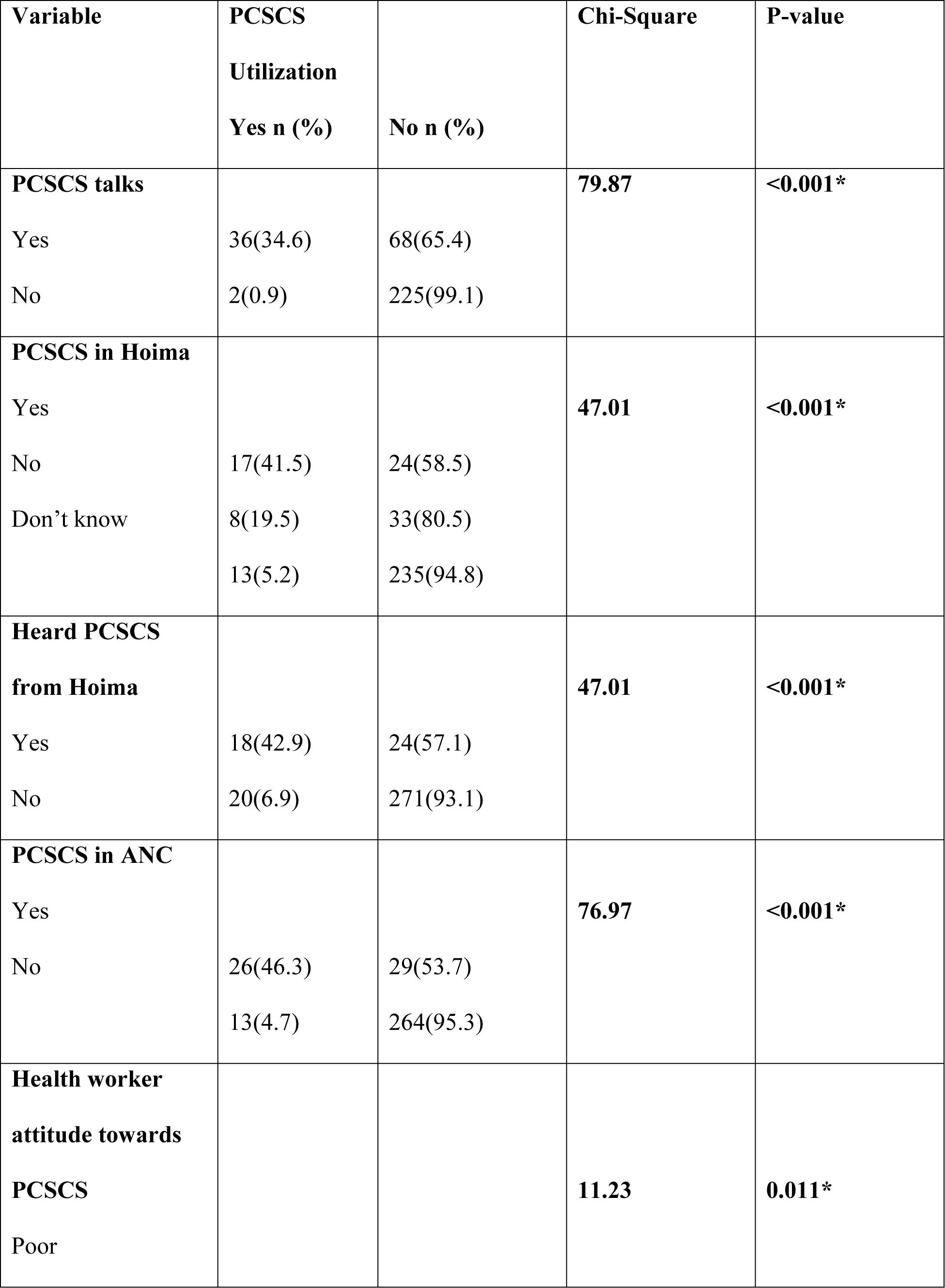
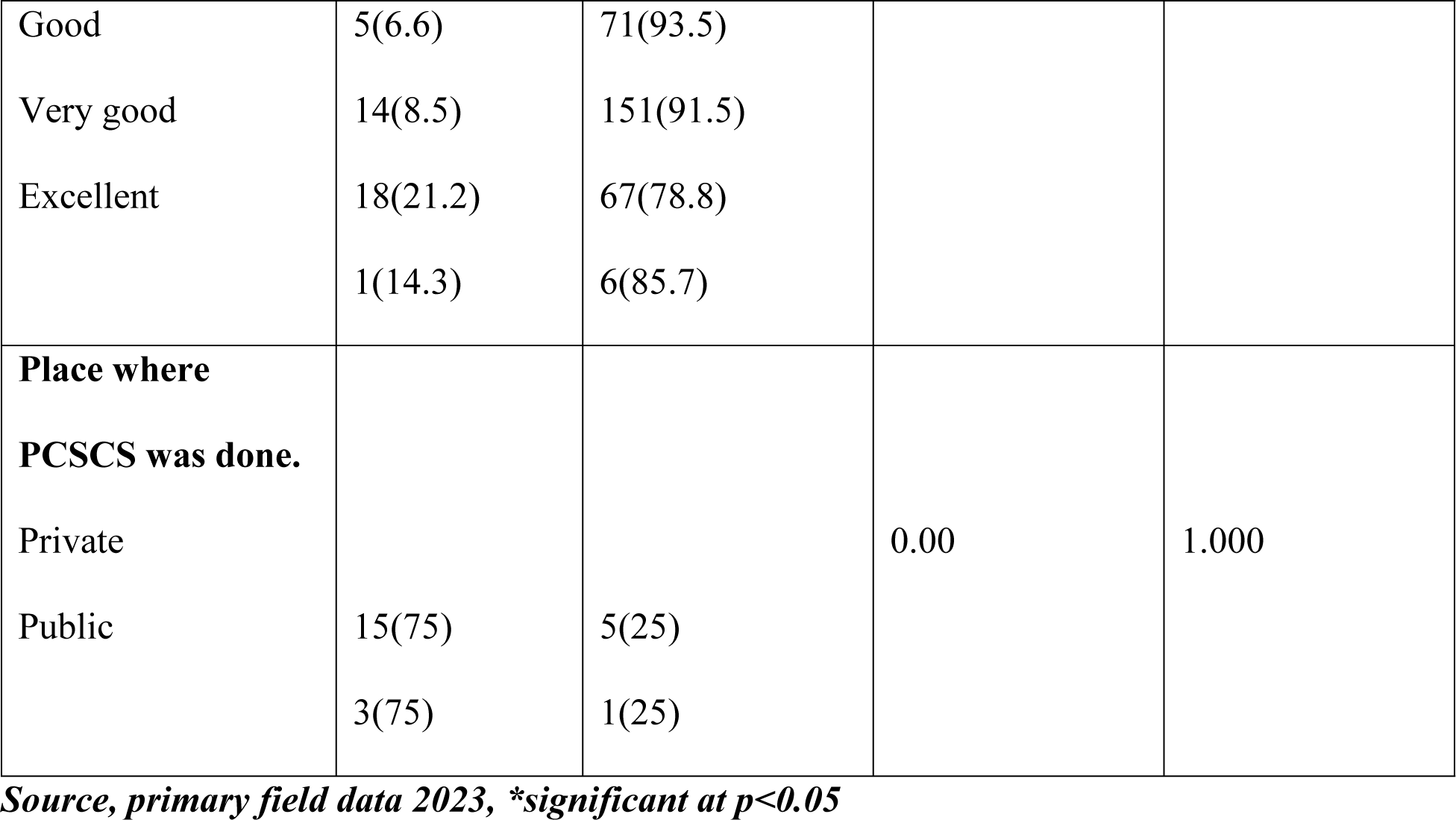
Bivariate analysis of Institutional factors associated with utilization of PCSCS among women 18-49 years receiving ANC from HRRH.

From the study, it was noted that women who had talks about PCSCS from different sources, radio, Television, church, health facilities or family members and friends utilized PCSCS 36(34.6%) more than those that didn’t have any talks in regards to PCSCS 2(0.9%). And for P-0.001, there was a significant relationship between PCSCS talks and utilization of PCSCS among women receiving ANC from Hoima RRH.

Secondly, women who were aware of having PCSCS at Hoima RRH 17(41.5%), utilized PCSCS more compared to those who didn’t know 8(19.5%) and those who were not sure of the presence of PCSCS at Hoima RRH 13(5.2%). For P-0.037, there was significant relationship between having PCSCS at Hoima RRH and utilization of the PCSCS among women attending ANC at Hoima regional referral hospital.

Thirdly, women who had heard about preconception sickle cell screening from Hoima RRH utilized PCSCS 18(42.9%) more than those who had never heard of it from Hoima RRH 20(6.9%). Since P-value of 0.001, there was significant association between having heard preconception sickle cell screening and utilization of PCSCS among women receiving ANC from HRRH.

Women who were solely receiving ANC services from Hoima RRH utilized PCSCS 26(46.3%) more than those who were not solely receiving ANC from Hoima RRH. And for P-0.001, there was a significant association between solely receiving ANC from Hoima RRH and utilization of PCSCS among women receiving ANC from Hoima RRH.

Lastly utilization of PCSCS was highest among those women who reported that the health workers attitude towards PCSCS was very good 18(21.2%), followed by those who reported the attitude was excellent 1(14.3%), those who reported being good 14(8.5%) and least among those who reported that the attitude was poor 5(6.6%). The P –0.011 there was a significant relationship between health workers attitude and utilisation of PCSCS among women receiving ANC from HRRH.

**Table 6:**
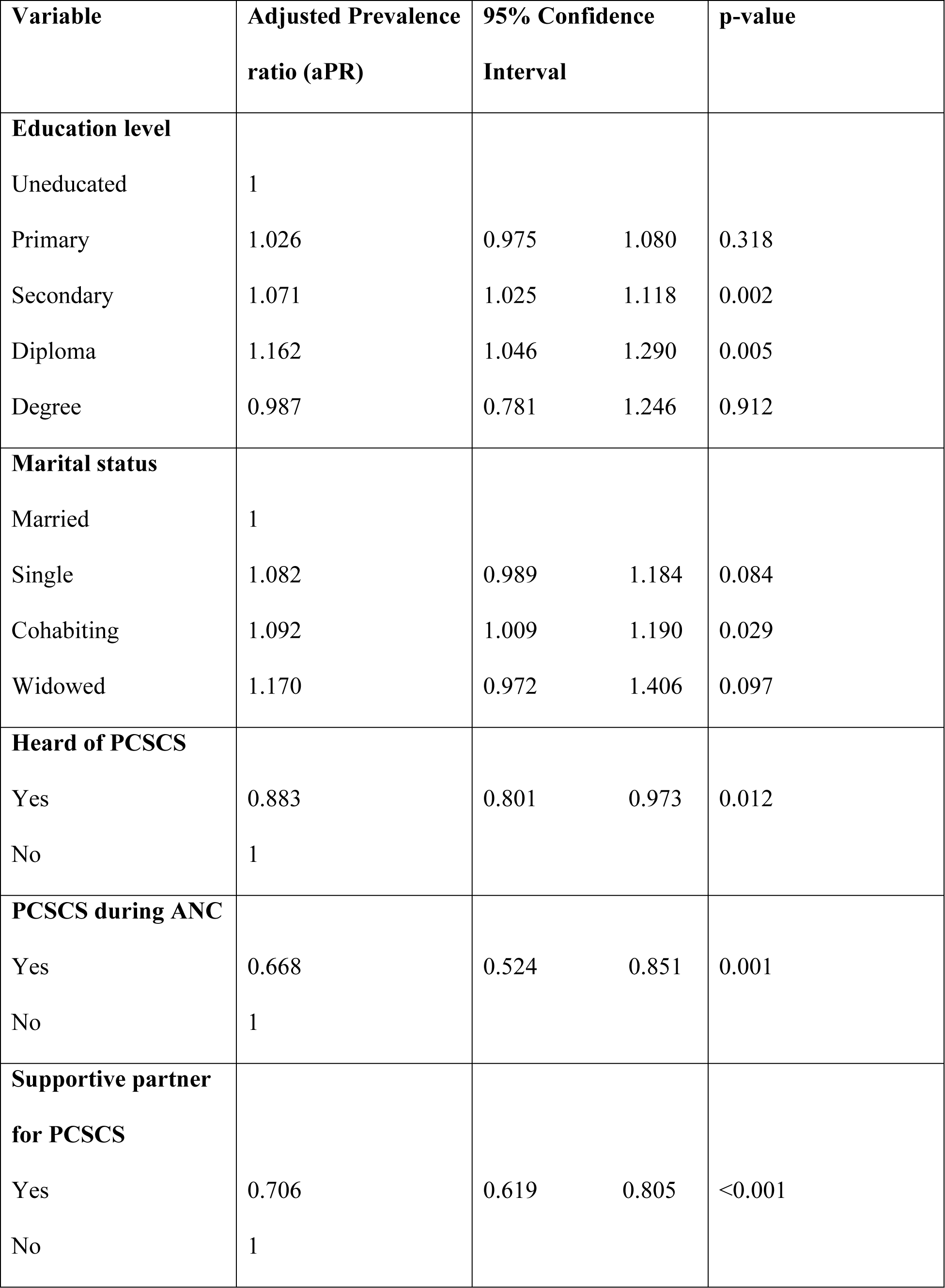
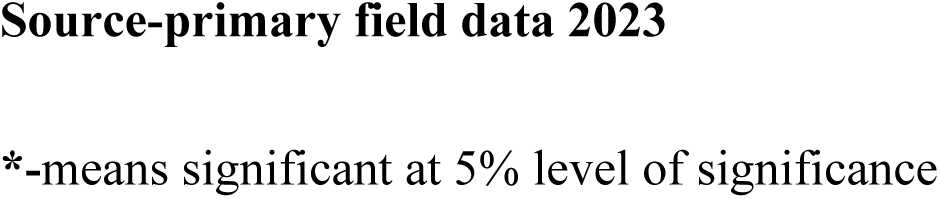
Multivariate analysis of variables of women 18-49 years with significant association with utilization of PCSCS.

| **Variable** | **Adjusted Prevalence**<br>**ratio (aPR)** | **95% Confidence**<br>**Interval** |  | **p-value** |
| --- | --- | --- | --- | --- |
| **Education level** |  |  |  |  |
| Uneducated | 1 |  |  |  |
| Primary | 1.026 | 0.975 | 1.080 | 0.318 |
| Secondary | 1.071 | 1.025 | 1.118 | 0.002 |
| Diploma | 1.162 | 1.046 | 1.290 | 0.005 |
| Degree | 0.987 | 0.781 | 1.246 | 0.912 |
| **Marital status** |  |  |  |  |
| Married | 1 |  |  |  |
| Single | 1.082 | 0.989 | 1.184 | 0.084 |
| Cohabiting | 1.092 | 1.009 | 1.190 | 0.029 |
| Widowed | 1.170 | 0.972 | 1.406 | 0.097 |
| **Heard of PCSCS** |  |  |  |  |
| Yes | 0.883 | 0.801 | 0.973 | 0.012 |
| No | 1 |  |  |  |
| **PCSCS during ANC** |  |  |  |  |
| Yes | 0.668 | 0.524 | 0.851 | 0.001 |
| No | 1 |  |  |  |
| **Supportive partner**<br>**for PCSCS** |  |  |  |  |
| Yes | 0.706 | 0.619 | 0.805 | <0.001 |
| No | 1 |  |  |  |

## Multivariate analysis done using Modified Poisson regression

Women who completed secondary school had 7.1% higher Prevalence of utilizing PCSCS than those who were uneducated (aPR=1.071, 95%CI: 1.025-1.118, P-0.002), while those who had completed tertiary had 16.2% higher prevalence of utilizing PCSCS than those who were uneducated (aPR=1.162, 95%CI: 1.046-1.290, P-0.005). Women who were cohabiting had 9.2% higher prevalence of utilizing PCSCS than those who were married (aPR=1.092, 95%CI 1.009-1.190, P-0.029).

Secondly, women who had ever heard of PCSCS had a 11.7% lower prevalence of utilizing PCSCS than those who had never heard of PCSCS(aPR=0.668,95% CI:0.801-0.973,P-0.012) while those who attended ANC only from Hoima had a 33.2% reduced prevalence of utilizing PCSCS compared to those who also attended ANC from other facilities(aPR=0.668,95%CI: 0.524-0.851,P-0.001)

Lastly women who received Partners’ support towards PCSCS had 29.4% reduced prevalence of utilizing PCSCS than those who didn’t receive any support (aPR=0.706, 95%CI: 0.619 –0.805, P-0.001).

## Discussion

The utilization rate of PCSCS among women attending ANC from Hoima regional referral hospital was found to be at 11.4%. This is not different from a similar study done in Mulago national referral hospital, located in the capital city, by Namukasa, 2015, that found the utilization rate of PCSCS at 11% among prim gravid mothers who were having their delivery services at the hospital. However the utilization rate is much lower compared to the global PCSCS utilization rate of 25% according to study done by Modell 2008. The utilization rate noted at Hoima regional referral hospital is slightly lower compared to that carried out in Nigeria 14% according to studies done by (4).

The utilization rate of PCSCS among women 18-49 years was lower compared to sickle cell screening rate among women in their reproductive ages found in the communities of Lubaga division, in capital Kampala that found the sickle cell screening rate at 19.6% in a study conducted by (Tusuubira at el, 2018) and also much lower compared to sickle cell screening rate among care givers of sickle cell children that were admitted in a rural hospital in northern Uganda, St Ambrosoli that was found at 63.5% by Ocen 2022, and also among university students at 24.4% by study done by Kisakye et al,2022. The utilization rate of 11.4% despite it being low, it was a bit higher to study done in Gabon in 2021, which found the utilization rate at 6% among 275 participants by study done by Mombo et al, 2021.

Therefore, the utilization of preconception sickle cell screening still remains low in both urban and rural settings as evidenced by the two studies in Hoima RRH, and Mulago national referral hospital.

Majority of respondents were aged between 18-35 years (98.7%), cohabiting women (42.3%), completed secondary education (49.6%) and 64% of the respondents were employed. Furthermore 64.7% of the respondents were living in urban areas, catholic 41.9%, 69.5% of them already had at least one child and 64.9% were Banyoro by tribe, 15.3% had familial history of sickle cell disease and 36.9% were earning between 100000-1million shillings. This was in line with study carried in the community of Lussajja a sub-urban area in Kampala with only difference that it involved both sexes, with 61.8% being females, majority being in age groups of 19-28 years 61.8%, 52.9% in marriage and 62.7% had biological children. In this same study 56.7% already knew their sickle cell status at the time of the study.

Also these respondents’ characteristics were similar to those of the participants for a study done in Mulago by Namukasa 2015, however it only considered only prim gravid mothers and of any age. The study differed from one done in Odisha by Bindhani et al, 2020 which included both male and female partners in the study, to determine the level of knowledge, attitude and utilization of preconception sickle cell screening. For a study done in Gabon by Mombo et al, 2021, 79% of the respondents who had preconception sickle cell screening were educated and were the age group of 16-28 years, however the specific level of education not stated.

The utilization rate of PCSCS was highest among women aged 18-35 years 11.9%, and least among those aged 36-49years 4.8%.there was no association between age of a woman and utilization of PCSCS. This was not different from the studies done elsewhere for example in a cross sectional study done in Saudi Arabia, Jazan region, it was noted that age was significantly associated with utilization of preconception sickle cell screening services and utilization was highest among women aged 25-30 years and lowest among those aged above 45 years (7). This was explained by study done by Mombo et al, 2021 which stated that women of a young age are highly knowledgeable and show good attitude towards utilization of PCSCS among women.

There was significant association between level of education and utilization of PCSCS among women 18-49 years receiving ANC from HRRH ( aPR=1.071,95%CI:1.025-1.118,P-0.002), and the utilization was highest among those who had completed university education23.1%, then tertiary13.3%, then secondary 10.9% primary 10.1% and none among the uneducated group. However the findings varied with some studies for example in a study done in Muhimbili hospital Tanzania there was a link between level of education and utilization of PCSCS. Utilization was highest among those who had completed secondary, then tertiary then university and there no direct relationship between level of education and utilization of PCSCS (Tusuubira et al., 2018. And this was not different from studies done in Oman by Alkalbani et al 2022a. this could however between noted by a review done by American society of hematology 2018, which reported that mothers level of education was independent of the mothers knowledge and awareness towards screening sickle cell in the preconception period.

There was no association between women employment status and utilization of PCSCS, however it was noted there was a higher utilization of PCSCS among the unemployed 15.3% compared to those who were employed 9.3%.These findings were different from those done before, for example study done in Mulago by Namukasa 2015, showed that mothers who were employed were more likely to have PCSCS done compared to those who are unemployed. The findings in Mulago were similar to those obtained by a study done in Benue city, Nigeria by Oluwole et al, 2022, that showed that unemployed women were 5.8 odds lower than employed women to utilize the PCSCS. This was explained by the availability of funds and easy access to both private and public facilities and easy access to commonly used media channels for PCSCS talks such as social media, televisions and radios. The reverse for Hoima could not be explained by this study and a further study on why it happened differently could be explained by a new study among the same population.

There was a significant association between marriage status and utilization of PCSCS among women receiving antenatal care from HRRH, at a 95% confidence interval during a multivariate analysis (aPR=1.092, 95%CI 1.009-1.190, P-0.029). The utilization of PCSCS was highest among those who were married 16.8%, cohabiting 9.2%, single 8.7% and none among the windows. This was due to the counseling sessions conducted among the partners in preparation for their marriage vows, in which church leaders would encourage them to have a couple screened for several diseases such as HIV, Hepatitis B, and sickle cell among them. The findings were similar to studies done in Saudi Arabia in which there was an association between marriage and utilization of preconception sickle cell screening, from the study it was noted that preconception sickle cell screening was made mandatory for all customary marriages (7). In another survey done in the two villages of Karaput district of Odisha, it was noted that married couples had high chances of utilizing PCSCS than the unmarried because they are highly knowledgeable, aware and 60% of them believe that PCSCS can prevent SCD and the utilization rate was 29% compared to the general population globally (Bindhani et al. 2020). From this study despite married ones having knowledge and screening for sickle cell, it was noted that in about 75% of those who screened it didn’t affect their choice of marriage. This was slightly lower to findings by the American society of hematology that found out that 88% of married who did the screening continued with their choices of marriage, but had early plans of having their children screened early to ascertain their sickle cell status.

For this study, there was no significant association between woman’s religious beliefs and utilization of PCSCS among women receiving ANC from Hoima regional referral hospital at 95% confidence intervals. However it was not that utilization of PCSCS was highest among Moslem 20.7%, followed by Anglicans 11.9%, then Pentecost 10.3%, catholic 10.0% and others by 0.0%. The findings were contrary to finds in study done in Cyrus and Sardinia, where preconception sickle cell screening was mandatory for all customary marriages and religion was considered one of the strategies to increase sensitization and knowledge of women about sickle cell disease (American Society of Hematology, 2018). Important to note, in Saudi Arabia despite making PCSCS mandatory for all the marriages, there was no evidenced reduction in SCD burden of the country since many of the couples continued to engage in producing children despite their SCD results (7).

From the study, there was a significant association between familial history of sickle cell disease and utilization of preconception sickle cell screening. It was noted that mothers who had a history of sickle cell disease utilized PCSCS 76.5% while those with no history utilized PCSCS 23.5%. This could be explained by the increased awareness and burden attributed by the disease due to prior experience. This similar to a study done in Mulago hospital by Namukasa 2015, where history of SCD was associated with utilization of PCSCS, mothers who had more than children utilized PCSCS more than the prim gravid, this was because more of them 65.4% had past experience and knowledge (Ocen and Ohurira,2022).

There was no association between mothers’ area of residence and utilization of PCSCS among mothers receiving ANC from HRRH, however it was noted that utilization was highest among those who resided in both areas 17.4%, then Urban 10.8% and least among the rural. This could be explained by the increased exposures to media talk shows in urban such as televisions, radios and availability of modern hospitals that provide PCSCS. This was not similar to study review by Osunkwo et al, 2021 in which areas of residence where highly associated with utilization of PCSCS among women and those in urban areas where more likely to have PCSCS done compared to those living in rural areas because of the closure distance to the facilities conducting the PCSCS services.

From the study there was no association between woman tribe and utilization of PCSCS, however the utilization of PCSCS was highest among Bakiga 15.0%, followed by Baganda 13.8%, then Banyoro 11.6%, others 9% and Bakonjo 0.0%. The findings were similar to study done among the caregivers of children with sickle cell who were receiving treatment from St Ambrosoli hospital, in this study there was no association between sickle cell screening and tribe or culture of the mother.

From this study, woman’s level of income was not associated with the utilization of PCSCS among women 18-49 years receiving antenatal care from Hoima regional referral. It was noted that however, that women who earned above 1000000 utilized PCSCS 83.3%. These were followed by those who didn’t earn anything 10.7% and then those who earned below 100000 10.0% and least by those who earned between 100000-1000000. The findings where contrary to a study done in the city of Lagos where woman level of income was associated with utilization of PCSCS among women in which those of a higher level of earning where more likely to utilize PCSCS compared to those of a lower level of earning. It was noted from the study that those of higher level, class 4 utilized PCSCS at rate of 49% (Oluwole et al, 2022). This was because the level 4 income mothers had funds to access the screening at both public and private facilities. This wasn’t noticed in the study at Hoima RRH, it was noted that those didn’t earn anything utilized PCSCS more than those who earned between 100000-1000000.

Mothers’ prior knowledge and awareness of PCSCS was significantly associated with utilization of PCSCS, and it was noted that mothers who were highly knowledgeable and aware of PCSCS utilized the screening 31.58% compared to those who were not aware of PCSC 0.9%. The findings were supported by the fact that the more one becomes aware and knowledgeable about the burden and associated complications the more one is likely to carry out the screening to allow early SCD screening and initiation on treatment incase couple decides to bare children. The findings were similar to those of a study done in Mulago hospital by Namukasa, 2015, that showed a positive association between mothers level of knowledge, awareness and utilization of preconception sickle cell screening services, from the study in Mulago, of the 11% who utilized PCSCS, 98% of them were much aware about the disease and 94% had a good understanding of the sickle cell disease and these findings were in agreement with the findings in a study done among care givers of children with sickle cell disease admitted in a hospital and from a community survey on sickle cell screening (Tusuubira et al, 2018). However this was not similar to study done in Nigeria that showed no significant relationship between mothers’ knowledge about PCSCS and its utilization. This was explained by despite mothers being knowledgeable, their choice to utilize PCSCS was highly affected by their attitude (17).

There was a significant association between partner support towards PCSCS and utilization among women receiving ANC from HRRH. It was noted from the study that those women who had support of their partners utilized PCSCS (39.1%) while those who didn’t receive support utilized PCSCS (1.9%). This is due because in cases where a male partner supports the female in ensuring the safety of the future child is well, women are more likely to comply and have the screening done since there is no associated resistance from the co –partner. This support can even be monetary to aid in paying for the test or even transport to reach the screening sites. The findings were to a study done in Tanzania, where there was a positive support of parents towards preconception sickle cell screening, thus providing support to ensure screening is done thus preventing seeing their children suffer with the burden related to sickle cell (Kisanga *et al.*, 2021).

From the study there was no significant association between distance traveled by mothers to HRRH and utilization of PCSCS. It was noted that utilization of PCSCS was highest among women who travelled distance above 5km 23.3%, travelled by those who travelled 1-5km 12.6% and least among those who travelled less than 1km 7.9% for ANC to HRRH. This was similar to study done among women who were caregivers of children admitted on wards at St Ambrosoli, there was no relationship between distance from the facility and utilization of sickle cell screening and testing services at the facility.

From the study, there was no significant association between cost or means of transport incurred or used by mothers to reach the facility. It was noted that utilization of PCSCS was highest among women who incurred costs above 10000 31.3%, followed by between 5000 –10000 19.2% and least among those who incurred less than 5000 9%. Majority of women travelled by Boda Boda about 89%, about 12% walked to the hospital and the rest used other modes of transport. This was similar to study done by Oluwole et al, 2022, the study didn’t find any association between means of transport or duration of travel to facility and utilization of PCSCS among women (Oluwole, et al, 2022).

From the study, there was a significant association between health talks about PCSCS among women 18-49 years receiving ANC from HRRH and utilization of PCSCS. The PCSCS was higher among those who had received health talks about PCSCS 34.6% than those who never attended these talks 0.9%. Mothers received talks from different sources such as radios, televisions, church counseling’s, family members and friends and also from health facilities.

The findings were similar to study conducted to assess knowledge, awareness and attitude towards sickle cell trait screening among university students in Uganda, conducted by (18). This was also similar to studies done in Saudi Arabia (7).

Additionally making PCSCS part of the ANC package talks to mothers showed a significant association with utilization of PCSCS, in same study it was noted that 46.3% had PCSCS during ANC. This was important to those that had missed during the prior preconception period and would utilize the service on the next preconception period or even plan to have their sickle cell test done during pregnancy to aid early childhood screening, diagnosis and treatment.

Availability of PCSCS was significant to its utilization among women receiving ANC from HRRH. Utilization of PCSCS was highest among mothers who were aware of the availability of PCSCS 41.5%, followed by those who didn’t know it was available 19.5% and then least among those who were not sure of its availability 5.2%. This was because mere knowing the service is available aided mothers in seeking for it compared to those who didn’t know or were not sure if it’s really available at Hoima RRH.

Findings were similar to those of a study carried out in Oman among 400 university students in which the availability of PCSCS increased its utilization rate and therefore was a significant relationship between availability of PCSCS and its utilization (Alkalbani et al., 2022).

In a cross sectional study carried out among 400 university students in Oman, despite these students being highly knowledgeable and aware of the availability of preconception SCD. Also these findings were similar to those in Uganda in a study carried out in Luuka district among women in their adolescent and reproductive age (15). However, these study findings were different to those in study done in the city of Lagos, Nigeria in which availability of preconception sickle screening services had no significant relationship with the utilization of services (Oluwole at el, 2022). These could have been affected by other factors such as cost of the service and women attitude in having the test done.

From the study done, there was no significant relationship between health workers attitude towards PCSCS and utilization of PCSCS among women 18-49 years receiving ANC from Hoima regional referral hospital. It was noted that utilization was high among women who reported the health workers attitude being very good 21.2%, followed by those who reported the attitude being excellent 14.3%, then good 8.55% and least among those who reported the attitude being poor 6.6%. This is contrary to what is expected as seen in a study done among caregivers where the health care workers attitude in having parents screened resulted in more caregivers screening for sickle cell disease.

From this study, there was no significant association between the cost of PCSCS and utilization of PCSCS among women attending ANC from HRRH. It is noted that the utilization of PCSCS was the same in both private and public health facilities at 75%. This contrary to study done by Alkalbani et al 2022b, to assess the willingness to undertake preconception sickle cell screening among women and in this study the cost of PCSCS was significantly associated with the utilization of PCSCS (19).

## Conclusion

The utilization rate of preconception sickle cell screening among women receiving ANC from Hoima regional referral hospital was low (11.4%). From the study, it was noted that the utilisation of PCSCS was significantly associated with woman’s level of education, marital status and support from partner towards PCSCS.

## Limitations to the study

There was a possibility of recall bias because women might not have been able to remember if they tested before conception, since it could have happened some time back and since laboratory evidence was not checked to a certain evidence of the screening, participants would more like reply depending on their desirability. This affected the internal validity of the study. In order to minimize this error, PCSCS was considered for that current pregnancy.

Participants declined to participate because topic was sensitive.

The study did not involve interviewing of both male and female and the information regarding male screening was obtained from their wives and the institutional related factors would have also been better understood by involving health workers as participants into the study.

Lastly, utilisation of PCSCS was also interfered with woman’s awareness of PCSCS and attending antenatal care only from HRRH.

## Recommendations

At individual level, local leaders need to empower women to attain a formal education through adult literacy program since a few of the participants were highly educated, majority had completed primary and secondary education.

At community level, DHOs office should ensure information in regard to PCSCS is translated into the native local languages. There is need for massive sensitization in community to enhance PCSCS knowledge on its benefits and also encourage women to attend ANC services through the VHTs.

At hospital level (HRRH), there is need by health workers to conduct regular health talks to women to enhance their level of knowledge and awareness and also ensuring the PCSCS is readily available.

## CRediT authorship contribution statement

Solomon Kyakuha^1^, Andrew Twineamatsiko^1*^: Project administration, Supervision, Investigation, Methodology & implementation.

Nathan Mugenyi^2^: Manuscript Writing – original draft, Writing – review & editing

## Declaration of competing interest

The authors declare that they have no known competing financial interests or personal relationships that could have appeared to influence the work reported in this paper.

## Data Availability

All relevant data are within the manuscript and its Supporting Information files.

## Acknowledgments

None

## Funding

None

## Notes

### Competing Interest Statement

The authors have declared no competing interest.

### Funding Statement

The author(s) received no specific funding for this work.

### Author Declarations

Mildmay Uganda Research Ethics Committee

